# Efficacy and Safety of Traditional Chinese Medicine in Obesity Management: A Systematic Review and Meta-Analysis

**DOI:** 10.64898/2026.06.04.26354905

**Authors:** Yanyan Wang, Yan Zhang

## Abstract

**Background:** Obesity is a global health crisis, contributing to chronic diseases such as diabetes, cardiovascular disease, and metabolic syndrome. Traditional Chinese Medicine (TCM) has been used in East Asia to manage obesity, but evidence on its efficacy and safety remains limited. This systematic review and meta-analysis assess clinical evidence from randomized controlled trials (RCTs) on TCM for obesity treatment.

**Methods:** We systematically searched PubMed, EMBASE, Cochrane Library, and Web of Science from inception to April 2026. Eligible RCTs compared TCM interventions with placebo or conventional treatments in obese patients. Two reviewers independently conducted screening, data extraction, and quality assessment. Meta-analysis was conducted using a random-effects model to calculate pooled weighted mean differences (WMD) and odds ratios (OR) for body weight, BMI, waist-to-hip ratio (WHR), lipid profiles, and adverse events.

**Results:** A total of 33 randomized controlled trials (RCTs) involving 3,053 participants were included in the analysis. TCM significantly reduced body weight (WMD = -5.86 kg, 95% CI: -7.51 to -4.21), BMI (WMD = -2.82 kg/m², 95% CI: -3.38 to -2.25), and WHR (WMD = -0.04, 95% CI: -0.06 to -0.02). Lipid profiles improved, with reductions in total cholesterol (WMD = -0.82 mmol/L), triglycerides (WMD = -0.65 mmol/L), LDL-C (WMD = -0.39 mmol/L), and increased HDL-C (WMD = 0.29 mmol/L) (all *p* < 0.001). Adverse events were infrequent, with no significant difference observed between TCM and control groups (OR = 0.51, 95% CI: 0.24 to 1.08). Funnel plots indicated no publication bias.

**Conclusion:** TCM appears effective in reducing body weight and improving lipid profiles in obese patients, with a low incidence of adverse events. It may serve as a complementary treatment for obesity, though further high-quality RCTs are needed to confirm these findings and assess long-term outcomes.

## Introduction

Obesity represents a significant global public health crisis, with prevalence rising rapidly in both developed and developing nations [1]. Currently, an estimated 2 billion adults worldwide are classified as overweight, of whom over 1 billion are obese [2]. This epidemic affects individuals of all ages, significantly burdening healthcare systems and economies globally. For instance, in the United States, approximately one-third of adults are obese, and in China, the combined prevalence of overweight and obesity in urban populations, particularly among children, has reached alarming levels [3]. Obesity is a major contributor to healthcare costs globally, with the U.S. alone spending an estimated USD 173 billion annually on obesity-related healthcare [4]. Obesity is a complex, multifactorial condition defined by excessive accumulation of body fat, which significantly increases the risk of numerous health complications and mortality [5]. The condition arises primarily from a prolonged positive energy balance, which is influenced by genetic predisposition, environmental factors, and behavioral aspects such as unhealthy dietary habits and insufficient physical activity [5]. The pathophysiology of obesity is characterized by metabolic dysregulation and chronic systemic inflammation, which exacerbate associated comorbidities [6]. Obesity is strongly linked to various serious health conditions, including cardiovascular disease, type 2 diabetes, hypertension, and metabolic syndrome [7]. Addressing this growing health crisis requires urgent action across multiple sectors.

Currently, obesity treatment approaches include lifestyle modifications, pharmacotherapy, and bariatric surgery, but each has significant limitations. Lifestyle interventions often struggle with poor long-term adherence and high relapse rates [8]. Pharmacological treatments may offer benefits but are frequently accompanied by side effects and demonstrate limited long-term efficacy [9]. Bariatric surgery, although effective for many individuals, is invasive and not universally suitable [10]. These challenges underscore the pressing need for complementary and alternative approaches that can manage obesity effectively and safely.

Traditional Chinese Medicine (TCM) offers a holistic approach to obesity management by addressing aspects such as energy regulation, blood circulation, and organ function [11]. TCM utilizes a variety of modalities, including herbal medicine, acupuncture, dietary therapy, and exercises like tai chi and qigong [12]. The mechanisms underlying TCM’s effects in obesity management are diverse, involving modulation of intestinal microflora, regulation of hormone levels, and alterations in fat metabolism [13]. Specific bioactive compounds derived from TCM, such as artemisinin, curcumin, celastrol, capsaicin, berberine, and ginsenosides, have shown potential in preclinical and clinical studies for managing obesity and related metabolic conditions [14]. In TCM, obesity is commonly conceptualized as a “dampness-heat” syndrome, a concept that modern biomedical research correlates with lipotoxicity, tissue hypoxia, and chronic low-grade inflammation, ultimately leading to insulin resistance and metabolic dysfunction [15]. Despite the historical and current utilization of TCM in obesity management, evidence supporting its efficacy and safety remains fragmented and often lacks the methodological rigor required for conclusive assessments. While individual studies and small-scale clinical trials have suggested promising results, these studies frequently face limitations such as small sample sizes, inadequate controls, and inconsistent outcome measures. There remains a critical gap in the consolidated, high-quality evidence required to assess the true effectiveness and safety of TCM interventions for obesity management. Specifically, there is an absence of systematic synthesis from randomized controlled trials (RCTs) that objectively evaluate the impact of TCM on obesity-related outcomes.

The aim of the present study is to conduct a systematic review and meta-analysis of RCTs to evaluate the efficacy and safety of TCM interventions in obesity management. By pooling data from multiple RCTs, this study seeks to provide an evidence-based understanding of TCM’s role in reducing body weight, improving BMI, WHR, and lipid profiles. Additionally, this review will examine the frequency and severity of adverse effects associated with TCM interventions to better delineate their safety profile.

## Methods

### Study Design

This study is a systematic review and meta-analysis designed to assess the efficacy and safety of TCM interventions in managing obesity. This study systematically identifies, evaluates, and synthesizes evidence from randomized controlled trials (RCTs) to assess the efficacy and safety of TCM interventions for obesity management. By combining data from multiple trials, this meta-analysis aims to provide robust and reliable conclusions regarding the impact of TCM on obesity-related outcomes, such as body weight, BMI, WHR, and lipid profiles, along with its safety profile. The study was conducted in accordance with the PRISMA (Preferred Reporting Items for Systematic Reviews and Meta-Analyses) guidelines, ensuring a systematic and transparent methodology [16].

### Search Strategy

A comprehensive literature search was conducted in eight databases: PubMed, EMBASE, Cochrane Library, Web of Science, China National Knowledge Infrastructure (CNKI), Wanfang Database, VIP Database, and SinoMed, from inception to April 2026. We also searched ClinicalTrials.gov and the Chinese Clinical Trial Registry (ChiCTR) for ongoing or unpublished trials. No language restrictions were applied. The protocol was registered with PROSPERO (CRD420261399439).

The search strategy employed a combination of keywords and Medical Subject Headings (MeSH) terms to identify pertinent studies. The search terms included: “Traditional Chinese Medicine”, “Herbal medicine”, “Acupuncture”, “Obesity”, “Body mass index”, “Waist-to-hip ratio”, “Lipid profile”, “Randomized controlled trials”, “RCTs”. These terms were used in various combinations to capture all relevant studies involving TCM interventions and their effects on obesity.

In addition to database searches, manual searches of reference lists from included studies were performed to identify any further relevant studies that may have been missed. Corresponding authors were also contacted when necessary to obtain unpublished data or to clarify study details.

### Eligibility Criteria

For this meta-analysis, only RCTs were included. Eligible studies needed to compare TCM interventions with a control group, which could involve placebo treatments, lifestyle interventions, or conventional treatments. Studies that lacked randomization or control groups, such as observational studies, case reports, or reviews, were excluded. Obesity was defined as BMI ≥ 30 kg/m^2^ (WHO criteria) or BMI ≥ 28 kg/m^2^ (Chinese criteria for Asian populations), in accordance with the criteria used in each original study, with no restrictions on participant age or geographic location to enable a comprehensive analysis of TCM’s effects globally. However, studies focusing exclusively on pediatric or non-obese populations were excluded. The interventions considered included various forms of TCM such as herbal medicine, acupuncture (both manual and electroacupuncture), and combination therapies like herbal medicine with acupuncture. Studies that involved alternative medicine approaches unrelated to TCM were not included. Comparators in eligible studies could include placebo treatments, pharmacotherapy, or lifestyle interventions such as diet and exercise, but studies that compared different TCM interventions without a control group were excluded. The primary outcomes evaluated in this meta-analysis were body weight reduction (measured in kilograms), improvements in BMI (kg/m²), and reductions in WHR. Secondary outcomes included improvements in lipid profiles, such as total cholesterol, LDL-C, HDL-C, and triglycerides, as well as any adverse reactions associated with the TCM interventions, like gastrointestinal symptoms or allergic reactions. Studies were excluded if they were not randomized controlled trials, lacked control groups, or were review articles, case reports, or non-human studies. Additionally, studies that did not focus specifically on obesity or failed to report primary outcomes, such as body weight or BMI changes, were excluded from the analysis.

### Data Extraction

Data extraction was performed independently by two reviewers. A pre-defined data extraction form was used to standardize the process and capture all relevant information from the included studies. The data items collected included the following: (1) study characteristics, such as author, publication year, and country of origin; (2) sample size; (3) details of the intervention, including the type of Traditional Chinese Medicine (herbal medicine, acupuncture, or combination therapies) and the duration of the treatment; (4) descriptions of the control group treatments (placebo, conventional treatment, or lifestyle intervention); and (5) the outcomes reported, with particular focus on primary outcomes such as body weight, BMI, and WHR, as well as secondary outcomes like lipid profiles and any reported adverse reactions.

### Risk of Bias Assessment

The quality of the included studies was assessed using the Cochrane Risk of Bias tool, which evaluates several domains to determine the potential for bias in randomized controlled trials [17]. These domains included: random sequence generation to ensure proper randomization, allocation concealment to prevent selection bias, blinding of participants and personnel to reduce performance bias, blinding of outcome assessors to control for detection bias, incomplete outcome data to address any missing data that could lead to attrition bias, selective reporting to detect reporting bias, and other potential sources of bias that could affect the validity of the study results. Each domain was rated as low risk, high risk, or unclear risk.

### Statistical Analysis

All data synthesis and statistical analyses were conducted using Review Manager (RevMan) and STATA software, which is widely used for meta-analyses in systematic reviews. For continuous outcomes, such as body weight, BMI, and lipid profiles, Weighted Mean Differences (WMD) were calculated to estimate the overall effect size. For dichotomous outcomes, such as the occurrence of adverse reactions, Odds Ratios (OR) were used. Both measures were reported with 95% confidence intervals (CIs) to provide estimates of precision. Heterogeneity among studies was assessed using the I^²^ statistic, which quantifies the percentage of variation across studies that is due to heterogeneity rather than chance. A value of I^²^ > 50% was considered indicative of substantial heterogeneity. To test the robustness of the results, sensitivity analyses were performed. Finally, publication bias was assessed through the use of funnel plots, which visually display the symmetry of the included studies’ effect sizes. In addition, Egger’s test or Begg’s test was used to statistically evaluate the likelihood of publication bias when appropriate, particularly in cases where visual inspection of the funnel plot suggested asymmetry.

### Ethics and Registration

As this study is a meta-analysis of previously published data, no ethical approval was required. All the data used in the analysis were extracted from studies that had already obtained ethical approval from their respective institutions.

## Results

### Study Selection

A total of 546 records were identified from the initial search of electronic databases, including PubMed, EMBASE, Cochrane Library, and Web of Science. After removing 259 duplicates, 259 unique studies were screened based on titles and abstracts. Of these, 224 records were excluded due to irrelevance to the topic, non-randomized designs, reviews, or lack of focus on obesity management. Subsequently, 35 full-text articles were assessed for eligibility, and 2 articles were further excluded (1 review article and 1 article not found). Ultimately, 33 studies were included in the qualitative synthesis, all of which were also included in the quantitative synthesis (Fig. 1).

**Figure 1.**
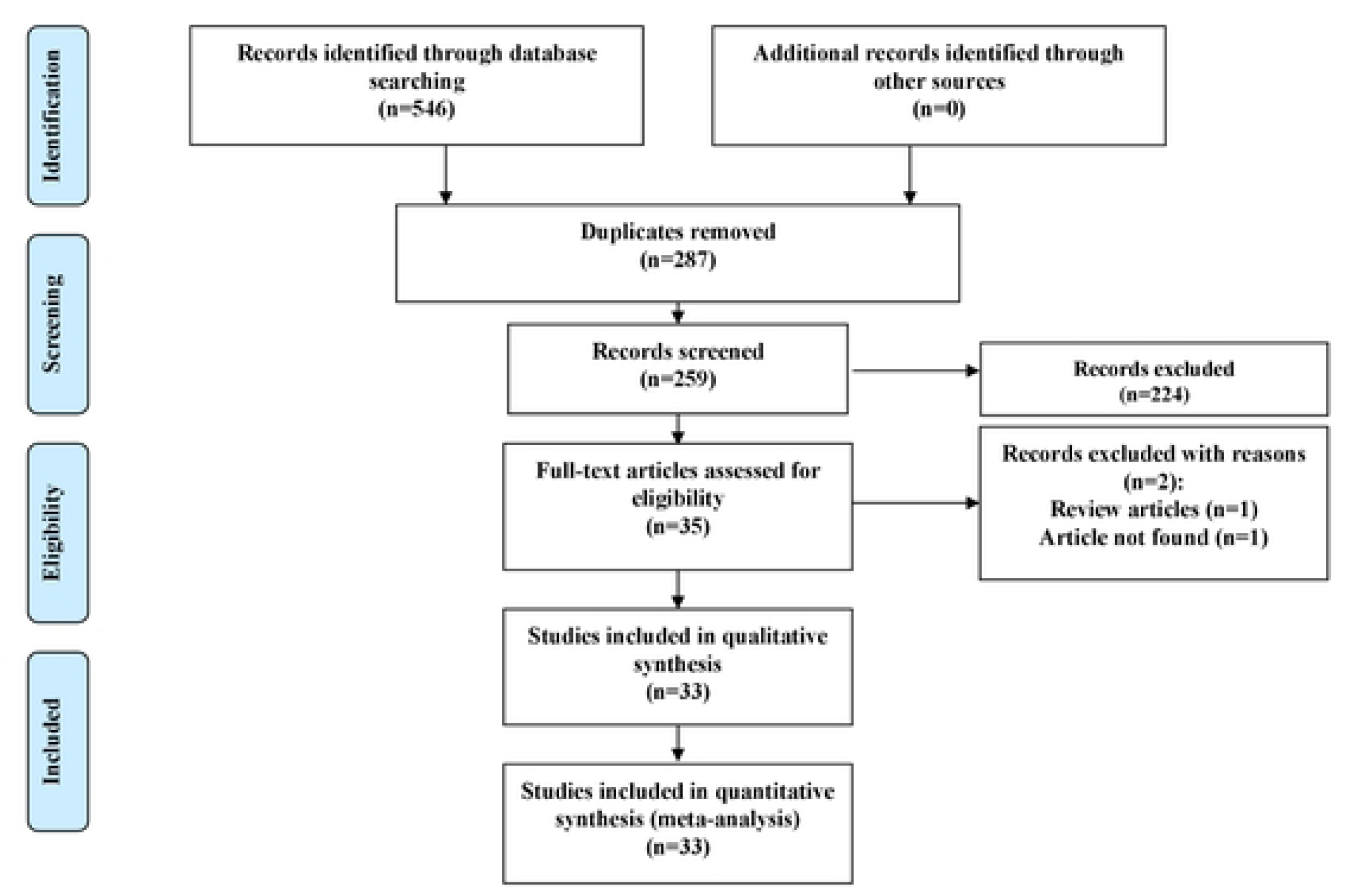
PRISMA Flow Diagram of Study Selection Process.

### Study Characteristics

The 33 included RCTs involved a total of 3,053 participants, with sample sizes ranging from 43 to 334 individuals per study. The studies were conducted in multiple regions, predominantly in China. The participants were classified as obese, with a BMI ≥ 30 kg/m² as the primary inclusion criterion across all studies.

Herbal medicine was the most common, utilized either as single herbs or in complex formulas. Acupuncture, including manual and electroacupuncture, was the second most frequently studied intervention. Combination therapies, incorporating both herbal medicine and acupuncture, were also included in several trials. The treatment duration varied across the studies, ranging from 8 to 24 weeks, with most studies following an 12-week treatment protocol. Control groups in the trials were treated with either a placebo, conventional treatments (such as weight-loss medications), or lifestyle modifications (e.g., diet and exercise programs). The primary outcomes reported in these studies were reductions in body weight, BMI, and changes in WHR. Secondary outcomes included improvements in lipid profiles, such as total cholesterol, LDL-C, HDL-C, and triglycerides, as well as the occurrence of adverse reactions related to TCM interventions. Detailed study characteristics and key outcome measures are summarized in Table 1.

**Table 1.** Basic Features of the Included Literature.

The risk of bias assessment shows that the majority of studies had a low risk of bias in key areas, including random sequence generation, allocation concealment, and selective reporting. However, there were some concerns related to blinding of participants and personnel (performance bias) and blinding of outcome assessment (detection bias), where some studies had unclear or high risk of bias. A small proportion of studies had a high risk of bias in areas such as incomplete outcome data (attrition bias) and other biases. Overall, the studies included in the analysis generally exhibited a low risk of bias across most domains, supporting the reliability of the meta-analysis findings (Fig. 2).

**Figure 2.**
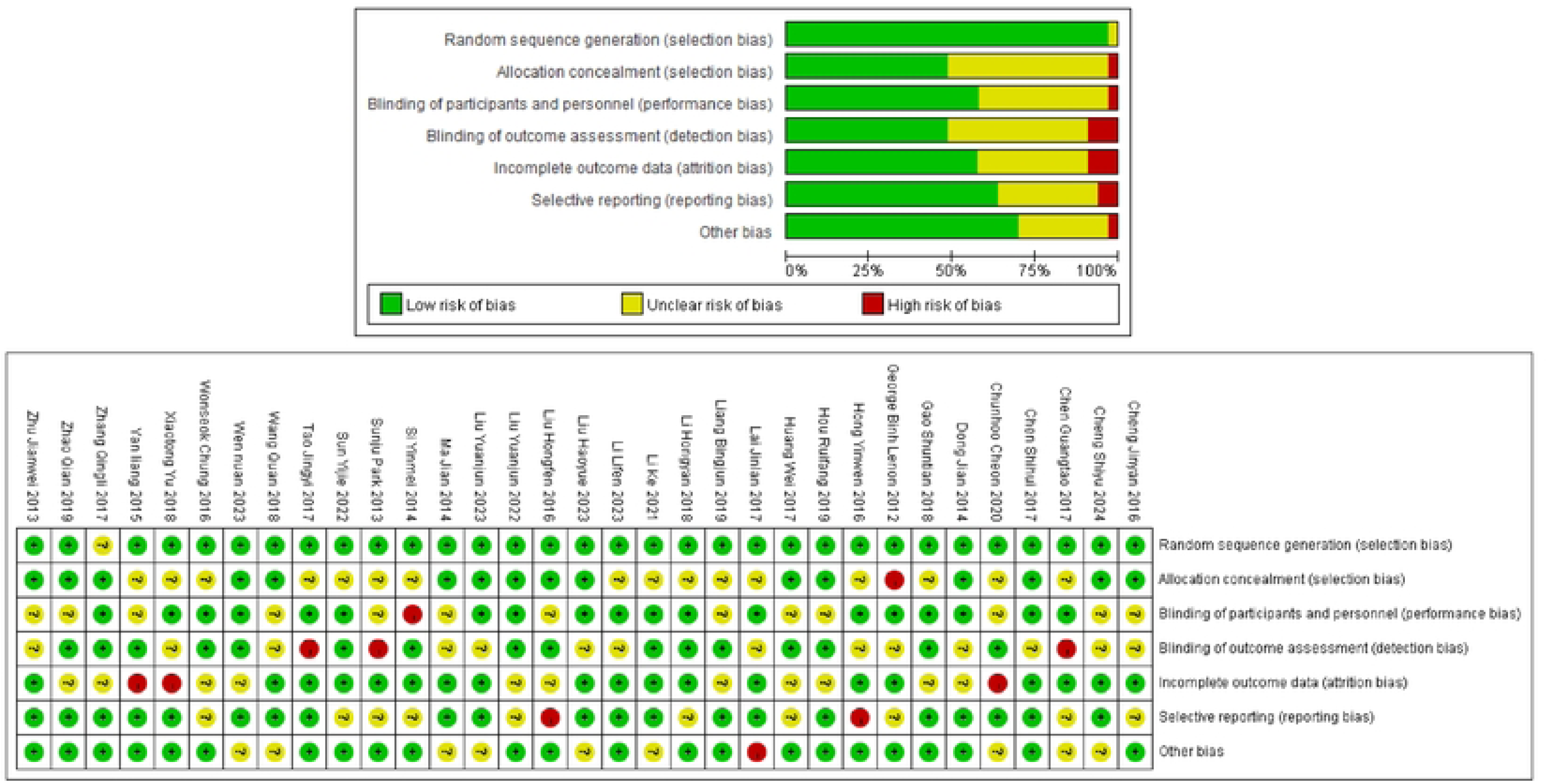
Risk of Bias Assessment.

### Primary Outcomes

The meta-analysis of the 33 studies showed that TCM interventions led to a significant reduction in body weight compared to control groups. The pooled WMD for body weight reduction was -5.86 kg (95% CI: -7.51 to -4.21, p < 0.001), indicating a substantial decrease in body weight among patients receiving TCM treatments. The analysis revealed considerable heterogeneity across the included studies (I² = 81.7%) (Fig. 3).

**Figure 3.**
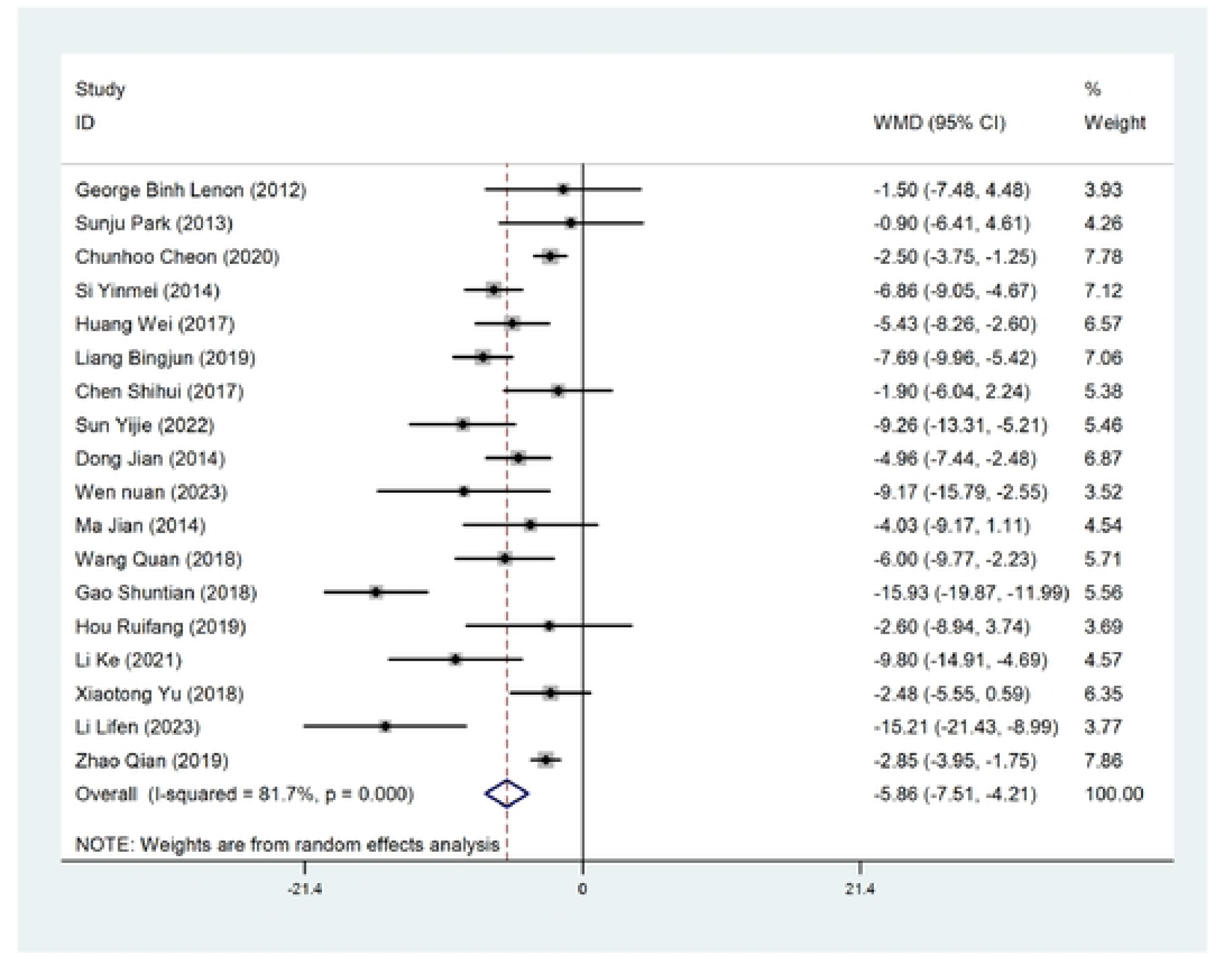
Forest Plot of Body Weight Reduction (kg) After TCM Treatment Compared to Control.

For BMI improvement, the meta-analysis demonstrated a significant reduction in BMI with TCM interventions compared to controls. The pooled WMD was -2.82 kg/m² (95% CI: -3.38 to -2.25, p < 0.001), indicating that TCM treatments effectively lowered BMI in obese patients. Heterogeneity was present among the studies (I² = 93.9%). The BMI improvement results were consistent across most studies, reinforcing the positive impact of TCM on body mass reduction (Fig. 4).

**Figure 4.**
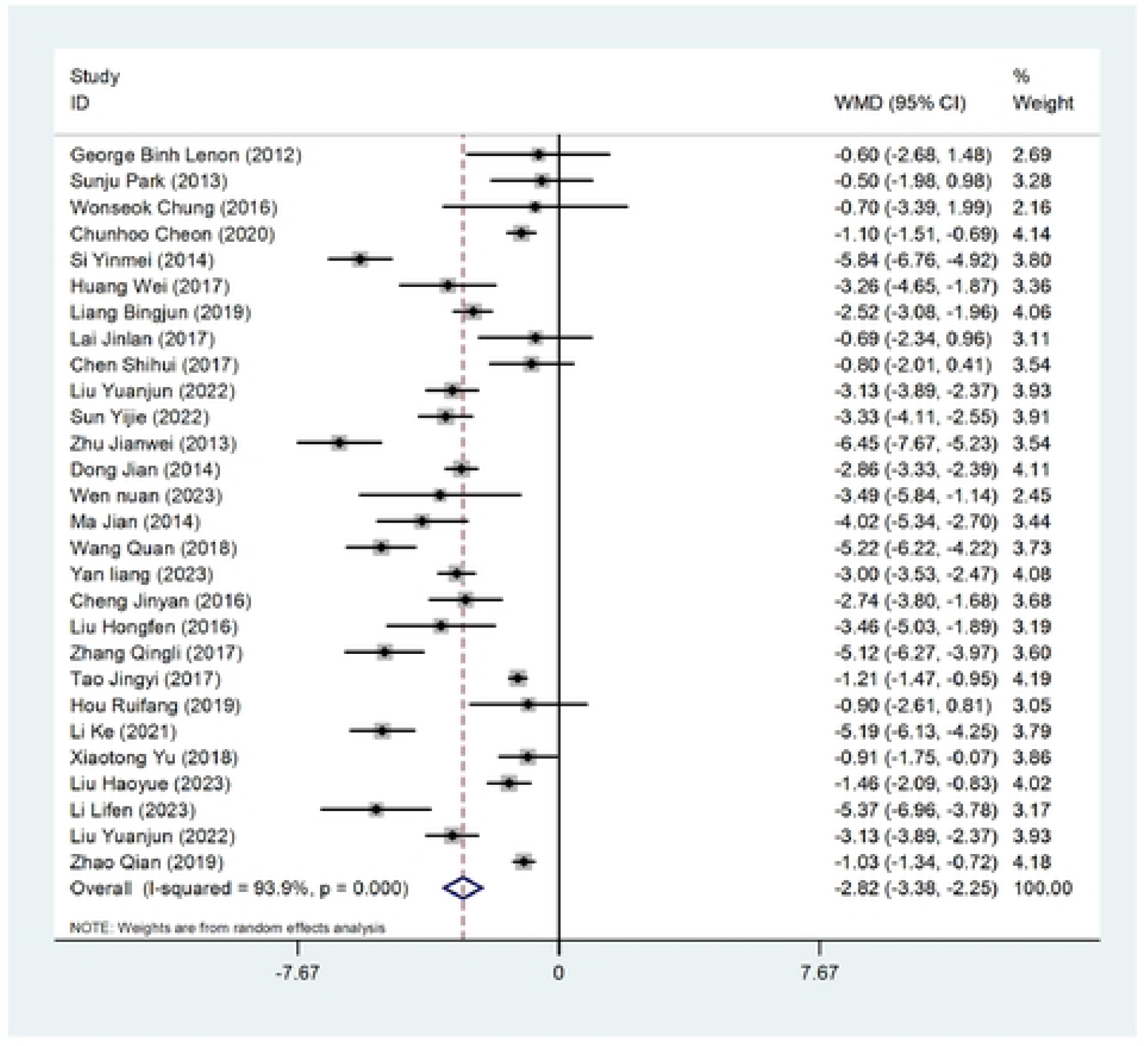
Forest Plot of BMI Reduction After TCM Treatment Compared to Control.

A significant reduction in WHR was observed in participants receiving TCM treatments. The pooled WMD for WHR reduction was -0.04 (95% CI: -0.06 to -0.02, p < 0.001). The heterogeneity for this outcome was relatively high (I² = 95.8%) (Fig. 5).

**Figure 5.**
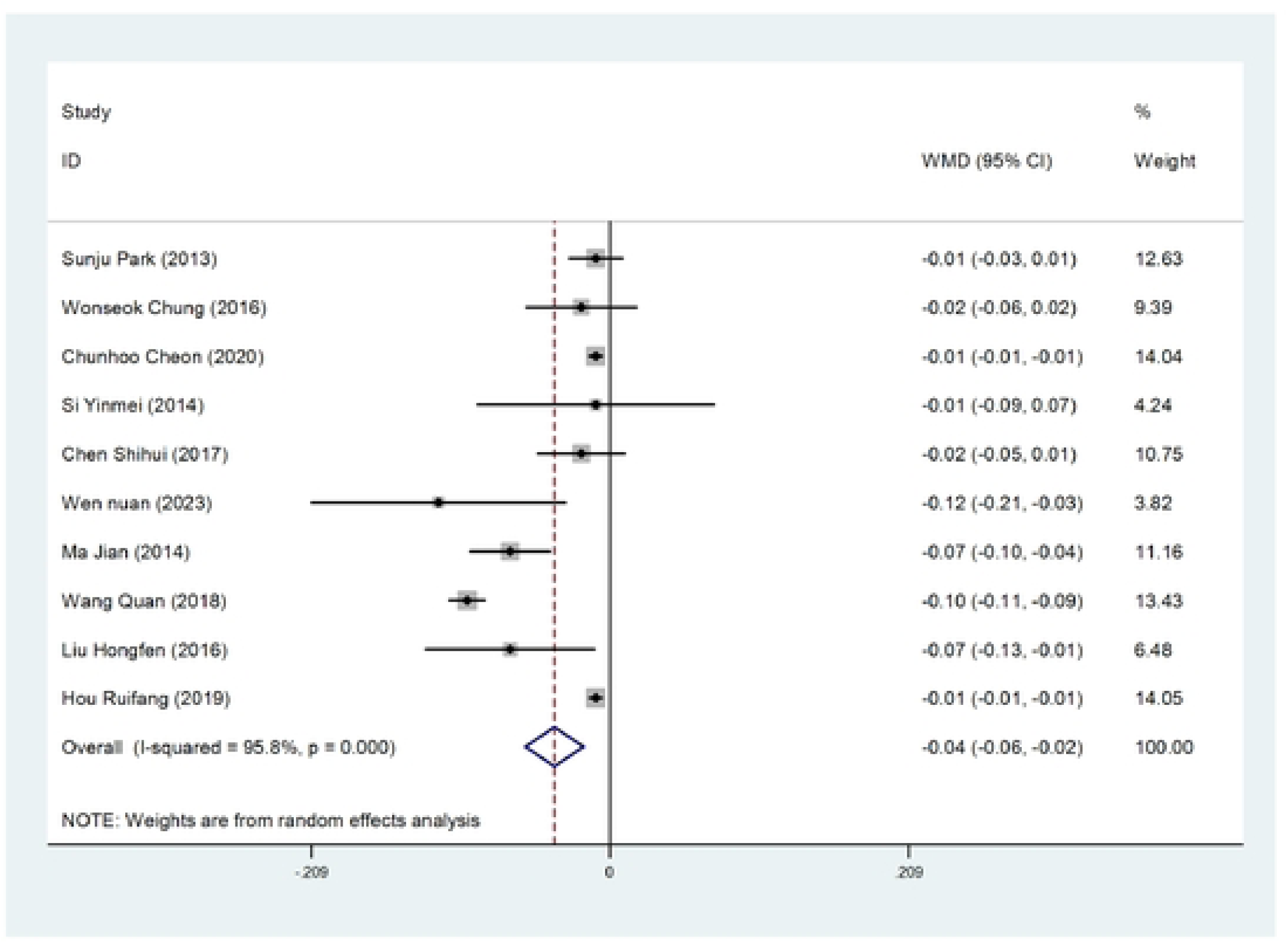
Forest Plot of Waist to Hip Ratio (WHR%) Reduction After TCM Treatment Compared to Control.

### Secondary Outcomes

The meta-analysis also evaluated the effects of TCM on lipid profiles, which are important markers of metabolic health. Significant improvements were observed in the following lipid parameters: The pooled WMD was -0.82 mmol/L (95% CI: -1.17 to - 0.46, p < 0.001), showing a marked decrease in total cholesterol levels among patients receiving TCM interventions (Fig. 6). A significant reduction in triglyceride levels was observed, with a WMD of -0.65 mmol/L (95% CI: -0.83 to -0.46, p < 0.001) (Fig. 7). The reduction in LDL-C was also significant, with a WMD of -0.39 mmol/L (95% CI: -0.59 to -0.19, p < 0.001) (Fig. 8). TCM treatments led to an increase in HDL-C, with a WMD of 0.29 mmol/L (95% CI: 0.14 to 0.44, p < 0.001) (Fig. 9). These results suggest that TCM interventions have a beneficial effect on lipid metabolism, improving cardiovascular health markers in obese patients.

**Figure 6.**
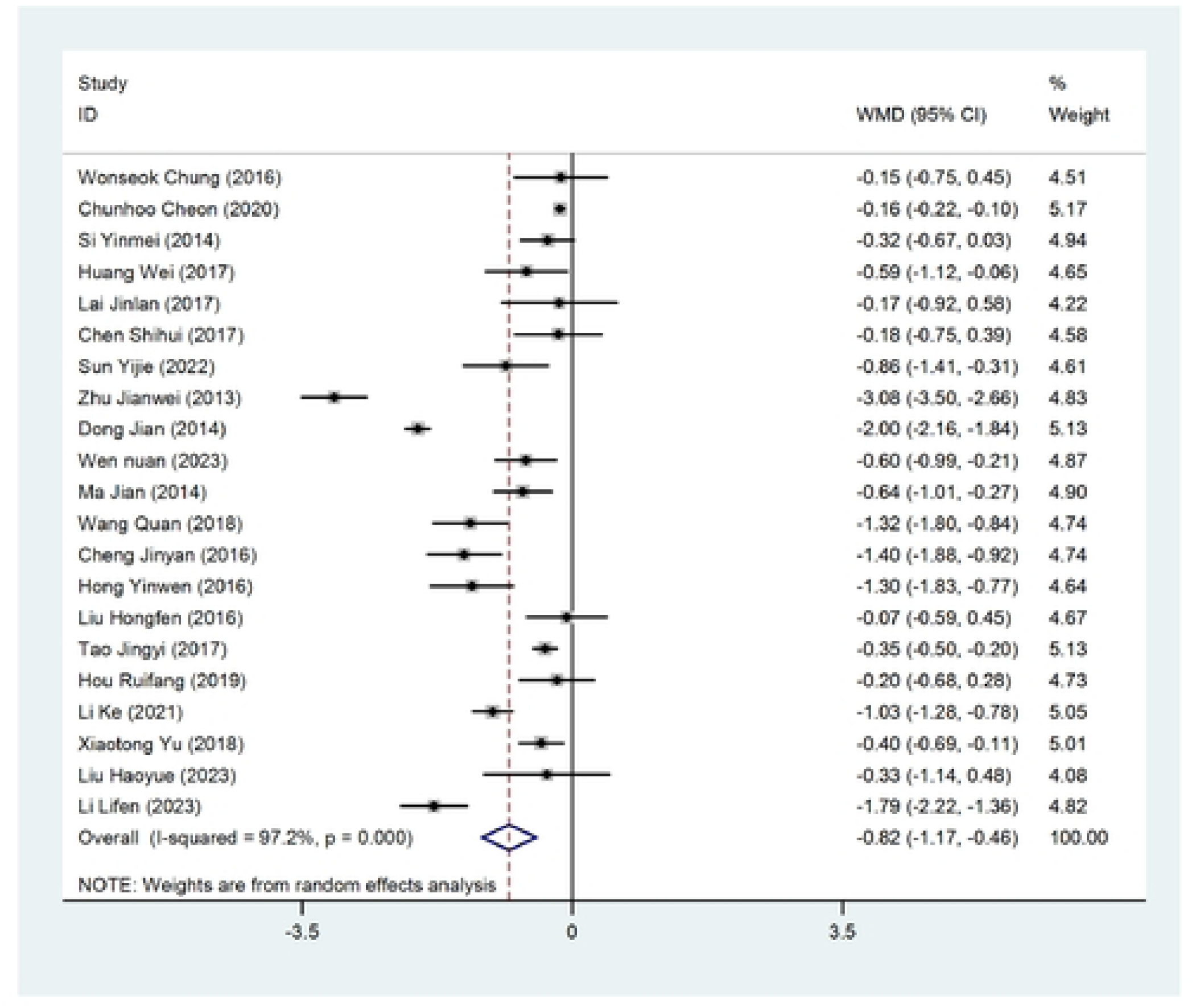
Forest Plot of Total Cholesterol (mmol/L) Reduction After TCM Treatment Compared to Control.

**Figure 7.**
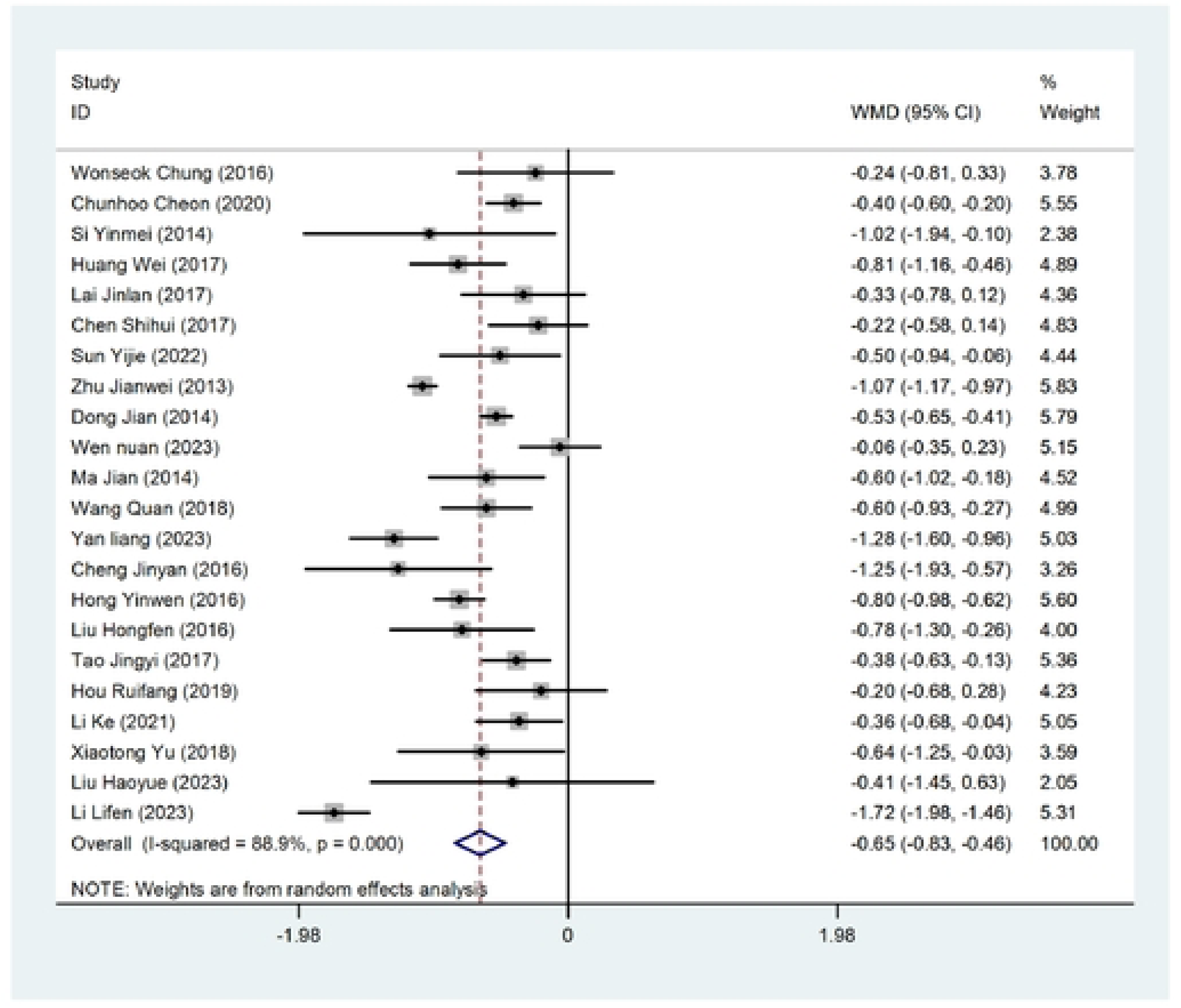
Forest Plot of Triglyceride (mmol/L) Reduction After TCM Treatment Compared to Control.

**Figure 8.**
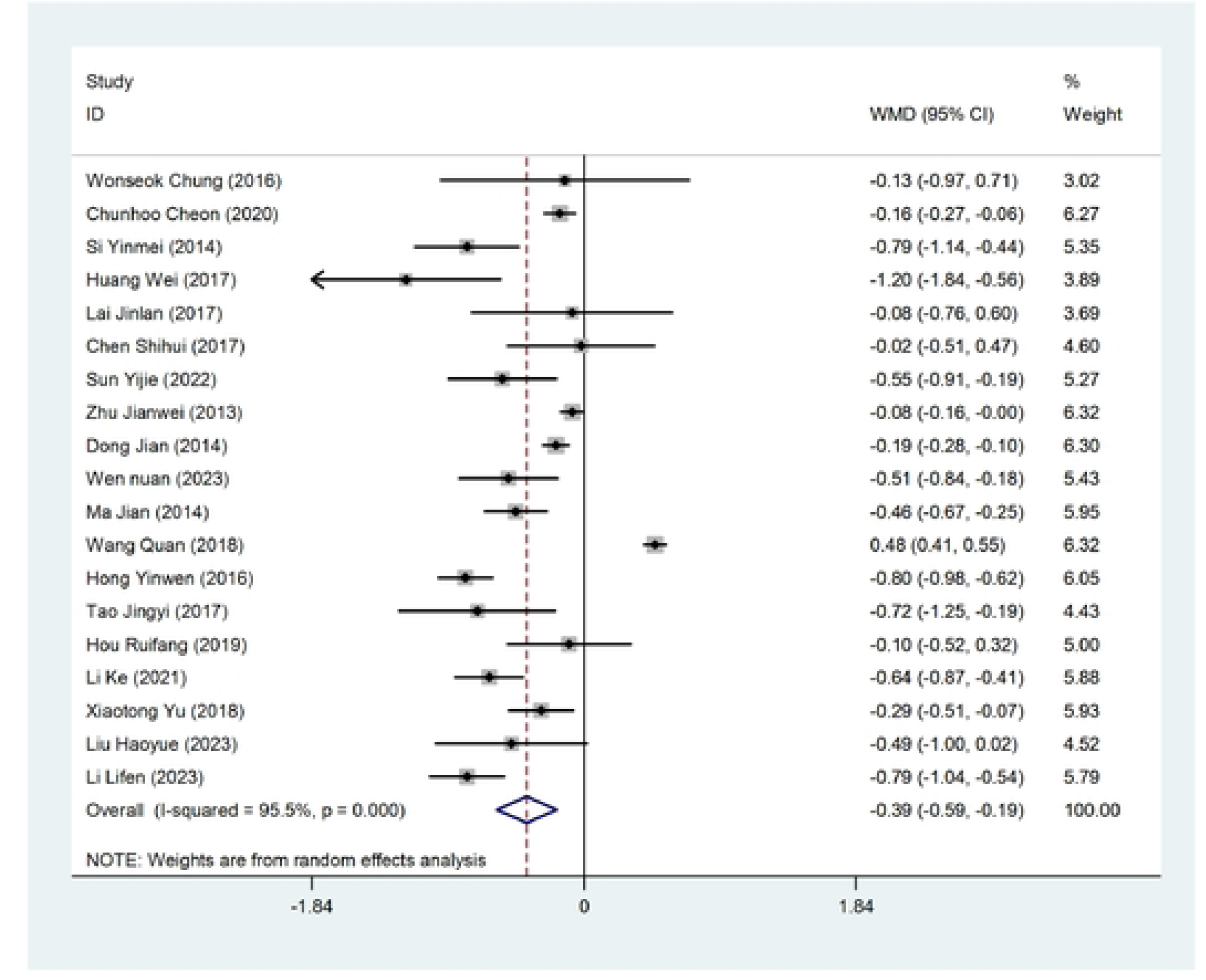
Forest Plot of LDL-C (mmol /L) Reduction After TCM Treatment Compared to Control.

**Figure 9.**
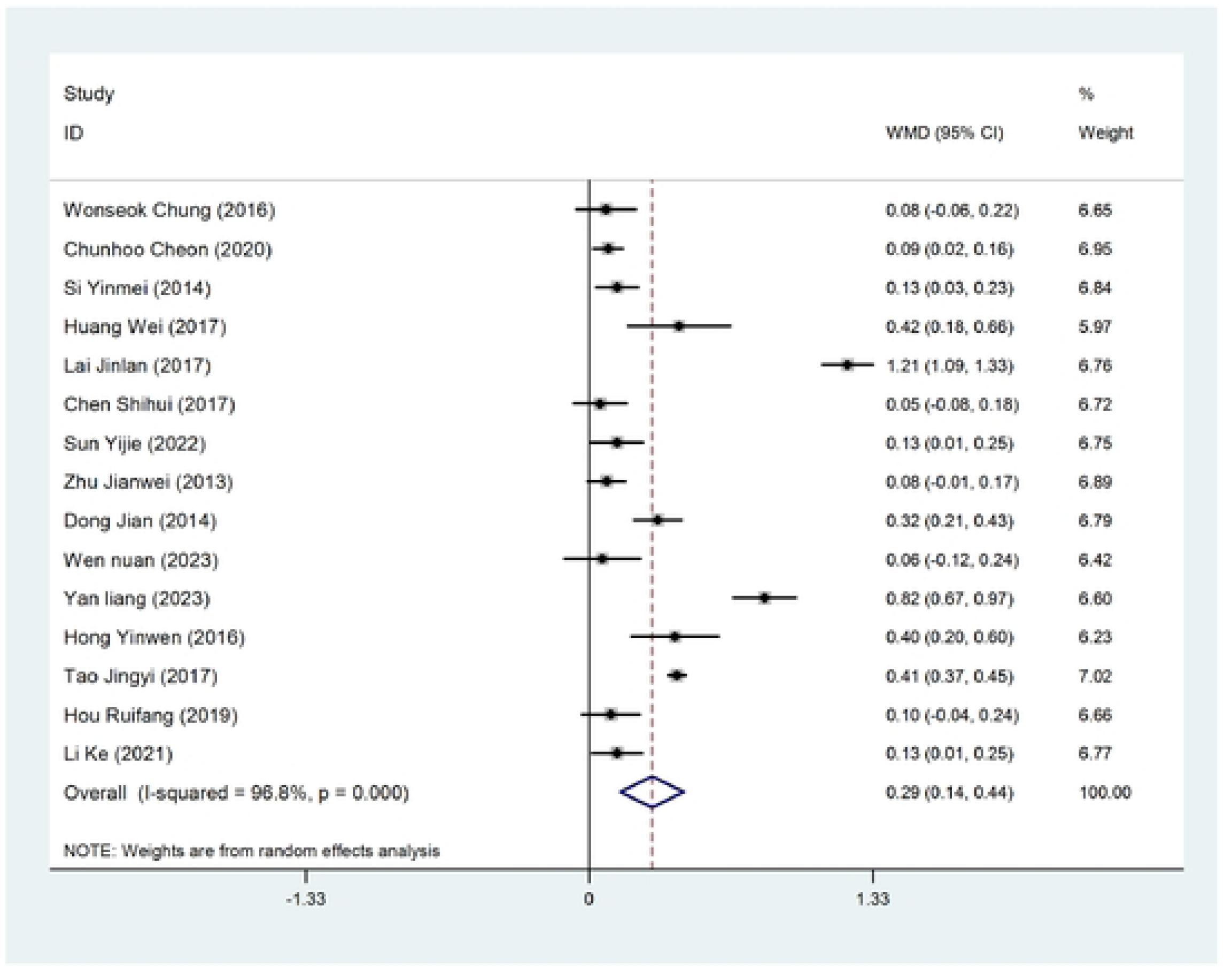
Forest Plot of HDL-C (mmol /L) Reduction After TCM Treatment Compared to Control.

### Adverse Reactions

A total of 9 studies reported data on adverse reactions related to TCM interventions. The pooled OR for the risk of adverse reactions in TCM-treated groups compared to controls was 0.51 (95% CI: 0.24 to 1.08) (Fig. 10), suggesting that TCM treatments were associated with a lower risk of adverse effects. The reported adverse reactions were generally mild and included gastrointestinal symptoms and allergic reactions. No serious adverse events were reported, further indicating the safety of TCM in the management of obesity.

**Figure 10.**
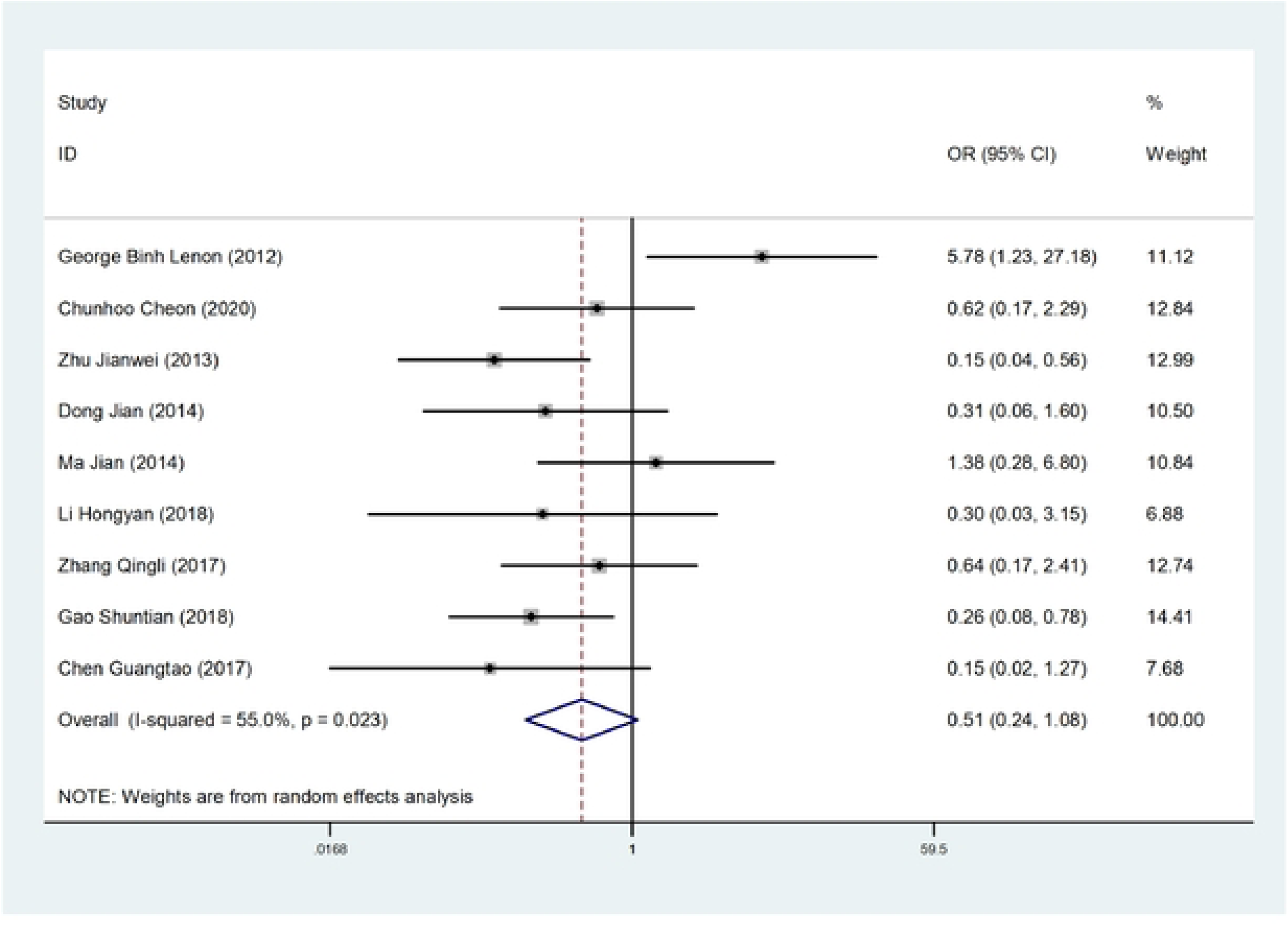
Forest Plot of Adverse Reactions After TCM Treatment Compared to Control.

### Funnel Plot and Sensitivity Analysis

To assess potential publication bias, funnel plots were generated for both the primary and secondary outcomes. Visual inspection of the funnel plots did not reveal any evident bias (Fig. S1). To evaluate the robustness of the meta-analysis results, a sensitivity analysis was conducted by systematically excluding individual studies to assess their impact on the overall effect size. The results of the sensitivity analysis indicated that the pooled estimates for body weight reduction and BMI improvement remained generally stable, with no single study having a disproportionate influence on the overall outcomes (Fig. S2). This demonstrates the robustness and reliability of the findings, despite the observed heterogeneity across studies.

## Discussion

The meta-analysis showed that TCM interventions, including herbal medicine, acupuncture, and combination therapies, were significantly more effective than control treatments (such as placebo, conventional therapy, or lifestyle interventions) in reducing body weight and BMI in obese patients. The pooled WMD highlighted the potential of TCM as a promising treatment for weight reduction. TCM interventions also demonstrated a notable impact on WHR, a key measure of central obesity, which is strongly associated with an increased risk of metabolic diseases such as cardiovascular disease and type 2 diabetes [18]. The meta-analysis showed that TCM significantly reduced WHR, highlighting its potential to address central obesity, a dangerous form of fat distribution linked to various health risks [19]. In addition to weight reduction, TCM interventions led to favorable changes in lipid profiles. Significant reductions were observed in total cholesterol, LDL-C, and triglycerides, while levels of HDL-C increased. These improvements in lipid profiles suggest that TCM not only helps with weight management but also promotes better cardiovascular health, potentially reducing the risk of heart disease and other metabolic disorders associated with obesity. The analysis of adverse reactions reported across the included studies indicated that TCM treatments were generally safe and well-tolerated. The pooled odds ratio showed a lower risk of adverse effects in patients receiving TCM compared to control groups. The reported side effects, such as mild gastrointestinal discomfort or allergic reactions, were infrequent and non-serious, further supporting the use of TCM as a safe intervention for obesity management.

### Comparison with Existing Literature

The findings of this meta-analysis align with previous studies and meta-analyses that have evaluated the role of TCM in obesity management. Acupuncture and related therapies, including electroacupuncture and acupoint catgut embedding, outperformed lifestyle modification alone in reducing body weight and BMI [20]. Combining acupuncture with other therapies ranked as the most effective method for weight and BMI reduction [20]. Acupuncture, both as a standalone treatment and in combination with lifestyle modifications, exhibited significant BMI reductions compared to control groups [21]. TCM treatments have shown promising potential for weight loss with fewer side effects, suggesting they could be considered as standalone or supplementary treatments for obesity in future clinical guidance.

However, there are some inconsistencies with prior research regarding the extent of lipid profile improvements. While our study found substantial improvements in total cholesterol, LDL-C, and HDL-C levels, not all previous meta-analyses reported such strong effects on lipid metabolism. For instance, a study found modest changes in lipid profiles, suggesting that the impact of TCM on lipid metabolism may vary depending on the type of intervention, the formulation of herbal treatments, or the patient population [22]. Our study’s inclusion of a wider variety of TCM interventions may explain the stronger lipid profile improvements observed here.

In terms of safety, the results of this meta-analysis align with existing evidence that TCM interventions are generally well-tolerated and pose a low risk of adverse effects. Previous trials and reviews have consistently reported that side effects associated with TCM treatments are mild and infrequent, which is consistent with our findings. In a review of Chinese herbal medicine for menopausal symptoms, adverse events were generally mild and rare [23]. A comparison of adverse drug reactions found that TCMs had lower occurrence frequency, severity, and hazard compared to Western medicines [24]. These findings consistently demonstrate the safety profile of TCM interventions across various conditions.

### Mechanisms of Action

The effectiveness of TCM in promoting weight loss and metabolic improvements can be attributed to several potential mechanisms. One of the primary ways in which TCM aids in weight reduction is through the regulation of lipid metabolism. TCM herbs can regulate lipid metabolism by influencing lipid absorption, synthesis, decomposition, and transportation [25]. Some herbal formulas enhance the activity of lipid-metabolizing enzymes like lipase, promoting the mobilization and utilization of fat stores [11]. Active ingredients from TCM, such as saponins, polysaccharides, alkaloids, and polyphenols, have demonstrated anti-obesity effects through various mechanisms, including appetite suppression, reduced lipid absorption, and increased lipid oxidation [26]. For instance, the Lingguizhugan decoction has been shown to improve fatty acid beta-oxidation and metabolism in high-fat-diet-induced fatty liver disease by modulating thyroid hormone levels and the expression of key genes involved in lipid metabolism [27]. These studies highlight the potential of TCM in addressing obesity and related metabolic disorders through multiple pathways.

Acupuncture has shown potential as a treatment for obesity by influencing appetite control and weight loss mechanisms. Studies suggest that acupuncture can affect the hypothalamus-pituitary axis, regulating hunger and satiety [28]. Acupuncture has been found to decrease body mass index, reduce eating desire and hunger, and increase gastric fluid survival rate compared to placebo treatments [29]. The therapy may work by increasing neural activity in the hypothalamus, enhancing stomach smooth muscle tone, and elevating levels of enkephalin, beta-endorphin, and serotonin [30]. These effects can suppress appetite, improve intestinal motility, and mobilize energy stores through lipolysis.

Obesity is characterized by excessive fat accumulation and is often associated with chronic low-grade inflammation, leading to metabolic disorders and insulin resistance. This chronic inflammation can impair the function of white and brown adipose tissues, exacerbating obesity and related conditions like diabetes and postprandial hyperlipidemia [31]. Traditional Chinese herbs, such as ginseng, have shown potential in treating obesity and diabetes through various mechanisms, including anti-inflammatory effects [32]. Additionally, some natural compounds like fucoxanthin have demonstrated anti-obesity and anti-diabetic effects by modulating adipokine levels and reducing inflammation in white adipose tissue [33].

### Strengths of the Study

This meta-analysis has several key strengths that enhance the reliability and validity of its findings. First, it included a large sample of 33 randomized controlled trials (RCTs), which increases the statistical power and enables more generalizable conclusions regarding the efficacy and safety of TCM interventions for managing obesity. Second, the study adhered to rigorous methodological standards, following PRISMA guidelines. This ensures a transparent, systematic process for study selection, data extraction, and quality assessment. Third, the analysis was comprehensive, assessing both efficacy and safety outcomes. In addition to primary outcomes like reductions in body weight, BMI, and WHR, the study also evaluated improvements in lipid profiles (total cholesterol, LDL-C, HDL-C, triglycerides) and monitored the incidence of adverse reactions. Finally, sensitivity analyses were employed to address potential heterogeneity, further strengthening the robustness of the findings by excluding studies with a high risk of bias or using alternative statistical models.

### Limitations of the Study

While this meta-analysis provides valuable insights into the efficacy and safety of TCM interventions for obesity management, several limitations need to be considered. One major limitation is the moderate heterogeneity observed in key outcomes, such as body weight and BMI reduction. This heterogeneity likely stems from variations in the study populations, differences in the types of TCM interventions used (e.g., herbal medicine, acupuncture, or combination therapies), and inconsistencies in treatment durations across the included studies. These factors introduce variability, making it more challenging to interpret the overall results with consistency. Another limitation concerns the quality of the included studies. Although the meta-analysis focused on RCTs, some studies exhibited a high risk of bias, particularly in areas like allocation concealment and blinding of participants and outcome assessors. These methodological shortcomings may have introduced bias, potentially impacting the reliability of certain results. The geographic concentration of the studies presents another constraint. The majority of trials were conducted in East Asia, particularly in China, where TCM is widely practiced. While this focus provides valuable insights into the effectiveness of TCM in populations familiar with its use, it raises concerns about external validity. The applicability of these findings to populations outside of East Asia remains uncertain, and further research in diverse geographic and cultural settings is necessary to determine the broader relevance of TCM interventions. Additionally, the potential for publication bias should be acknowledged. Although funnel plots and Egger’s test were used to assess this risk, it is still possible that smaller or negative studies were less likely to be published or included in the analysis, leading to an overestimation of TCM’s positive effects. While the tests did not strongly indicate publication bias, the possibility cannot be entirely ruled out.

### Future Research Directions

While this meta-analysis provides valuable insights into the efficacy and safety of TCM interventions for obesity management, several areas warrant further investigation. Future research should focus on addressing the current gaps in the literature to enhance our understanding of TCM’s long-term benefits and its broader applicability. One key area for future research is the long-term efficacy and safety of TCM interventions. While the included studies demonstrated significant short-term improvements in body weight, BMI, and lipid profiles, there is limited evidence on the sustained effects of TCM treatments over extended periods. Long-term follow-up studies are needed to assess whether the initial benefits of TCM are maintained and whether there are any delayed adverse effects or complications associated with prolonged use of TCM interventions. Additionally, there is a need for large-scale, high-quality RCTs conducted in diverse populations outside of East Asia. The current evidence base is heavily concentrated in studies conducted in China and other East Asian countries, where TCM is traditionally practiced. To generalize the findings to broader populations, future trials should be conducted in regions with different cultural, dietary, and lifestyle backgrounds. Another promising area for future investigation is the exploration of the specific bioactive components in TCM formulations that contribute to weight loss and metabolic regulation. While TCM herbal mixtures have shown efficacy, the precise mechanisms by which they promote weight loss are not fully understood. Research focusing on the pharmacological properties of individual herbs and their active compounds can elucidate how they regulate lipid metabolism, control appetite, and reduce inflammation. Comparative studies are also needed to evaluate the relative effectiveness of different types of TCM interventions, including herbal medicine, acupuncture, and combination therapies. Additionally, exploring the optimal combination of herbal and acupuncture therapies, including the appropriate dosage and frequency, could enhance the overall effectiveness of TCM treatments.

## Conclusion

This meta-analysis demonstrates that TCM interventions, including herbal medicine, acupuncture, and combination therapies, are effective in reducing body weight, BMI, and waist-to-hip ratio, as well as improving lipid profiles in obese patients. The findings suggest that TCM can serve as a valuable complementary therapy for obesity management, offering both metabolic benefits and a favorable safety profile. While the results are promising, further research is needed to confirm the long-term efficacy and safety of TCM, especially in diverse populations outside of East Asia.

## Data Availability

No new data were generated or analyzed during this study. All data used in this systematic review and meta-analysis are from previously published studies, which are cited and referenced in the manuscript.

## Authors’ contributions

Yanyan Wang designed the study. Yan Zhang analyzed the data, participated in the data collection, and prepared the manuscript. Yanyan Wang and Yan Zhang helped the analysis with constructive discussions. All authors critically revised the manuscript.

## Availability of data and materials

The original contributions presented in the study are included in the article/ supplementary material. Further inquiries can be directed to the corresponding authors.

## Declarations

Not applicable.

## Consent for publication

Not applicable.

## Figure legends

Figure S1. Funnel Plots for Assessing Publication Bias.

Figure S2. Sensitivity Analysis for the Primary and Secondary Outcomes.

